# The association between blast exposure and transdiagnostic health symptoms on systemic inflammation

**DOI:** 10.1101/2021.04.08.21255173

**Authors:** Jasmeet P. Hayes, Meghan E. Pierce, Kate E. Valerio, Mark Miller, Bertrand Russell Huber, Catherine B. Fortier, Jennifer R. Fonda, William Milberg, Regina McGlinchey

**Author notes:** **Address Correspondence to: Jasmeet P. Hayes, Ph.D.**, Department of Psychology, The Ohio State University, 225 Psychology Building, 1835 Neil Avenue, Columbus, OH 43210.

## Abstract

Chronic elevation of systemic inflammation is observed in a wide range of disorders including PTSD, depression, and traumatic brain injury, all of which are relatively common in United States Veterans. Although previous work has demonstrated a link between inflammation and various diagnoses separately, few studies have examined transdiagnostic symptoms and inflammation within the same model. The objective of this study was to examine relationships between psychiatric and health variables and systemic inflammation, and to determine whether mild traumatic brain injury (mTBI) and/or exposure to blast munitions moderate these relationships. Confirmatory factor analysis in a large sample (N = 357) of post-9/11 Veterans demonstrated good fit to a four-factor model reflecting traumatic stress, affective, somatic, and metabolic latent variables. Hierarchical regression models revealed that each of the latent variables were associated with higher levels of systemic inflammation. However, the strongest relationship with inflammation emerged among those who had both war-zone blast exposures and metabolic dysregulation, even after adjusting for mental health latent variables. Exploratory analyses showed that blast exposure was associated with metabolic dysregulation in a dose-response manner, with self-reported closer blast proximity associated with the greatest metabolic dysregulation. Together, these results provide greater understanding of the types of symptoms most strongly associated with inflammation, and underscore the importance of maintaining a healthy lifestyle to reduce the impact of obesity and other metabolic symptoms on future chronic disease in younger to middle-aged Veterans.

## INTRODUCTION

Systemic inflammation is observed in a wide spectrum of health conditions, from depression and posttraumatic stress disorder (PTSD), to obesity and hypertension [1]. Inflammation is a complex process that has both positive and negative effects on health. Although the acute action of pro-inflammatory cytokines such as interleukin-6 (IL-6) and tumor necrosis factor (TNF)-α are vital for the body’s response to infection, chronic elevation of these factors impede the body’s capacity for repair and is associated with diabetes [2], cognitive decline [3], and Alzheimer’s disease [4].

Considering that Veterans of the post 9/11 era commonly have high rates of comorbidity of mental health disorders, along with metabolic alterations and non-specific somatic symptoms including chronic pain [5], this population may be particularly vulnerable to the negative long-term health effects of chronic inflammation. PTSD and depression are associated with increased systemic inflammation in military personnel [6,7], as are conditions associated with metabolic dysregulation such as obesity [8], highlighting the transdiagnostic nature of symptoms associated with inflammation.

Further complicating the clinical picture in Veterans, the documented prevalence of blast exposure and/or blunt head impacts during deployment raises concern for vulnerability to disease secondary to inflammation. Among Veterans with traumatic brain injury (TBI), higher PTSD symptom severity is associated with elevated inflammation [9]. Recent data in animal models show that head injury, when combined with subsequent stress exposures, increases inflammation [10]. In this way, TBI may predispose the nervous system to be more vulnerable to subsequent CNS injuries through hyperactivation of microglial responses [11]. Blast exposure has also been associated with elevated inflammation; breakdown of the blood brain barrier [12] may facilitate penetration of circulating inflammatory cytokines that increase risk for dementia [13]. One-third of TBIs in military personnel originate from blast-related mechanisms such as improvised explosive devices and rocket propelled grenades [14], raising the concern that head injury and blast exposures modify and exacerbate psychiatric and medical comorbidities and their relationship with inflammation in humans.

The primary aim of this study was to examine associations between mental and metabolic health symptoms and inflammation, and further, the moderating effect of blast exposure and mild TBI (mTBI) on these relationships. Although previous work has shown relationships between inflammation and various diagnoses separately, few studies have examined the influence of transdiagnostic health symptoms on systemic inflammation in the same model. The last decade has seen a greater movement toward examining mental health symptoms dimensionally, in recognition of the heterogeneity of symptoms within and across categorical diagnoses [15–17]. We used confirmatory factor analysis (CFA) to examine the fit of transdiagnostic factors within a rich dataset from the Translational Center for TBI and Stress Related disorders (TRACTS) study [18]. We reasoned that examining health symptoms dimensionally would reveal whether some symptoms (e.g., somatic symptoms) would be more strongly associated with inflammation than others (e.g., affective). Metabolic health indicators (e.g., body mass index [BMI], waist-to-hip ratio, high-density lipoprotein [HDL]) were included in the model based on the well-established evidence indicating the role of these factors in inflammation [19]. Following the identification of factors from the CFA, each symptom factor was examined in relationship to pro-inflammatory cytokine concentrations and the moderating influence of mTBI and blast exposure. We capitalized on recent advances in fluid biomarker technology [20] to measure TNF-α and IL-6 using Simoa technology (Quanterix). In addition, C-reactive protein (CRP) was examined given the vast literature implicating elevation of this pro-inflammatory marker in various health conditions. We hypothesized that exposure to blast munitions and mTBI would interact with each of the latent variables to predict elevated inflammation.

## MATERIALS AND METHODS

### Participants

The final sample included 357 participants who were consecutively enrolled post-9/11 Veterans from the longitudinal TRACTS cohort study at VA Boston Healthcare System, a TBI National Network Research Center (see [18,21] for study recruitment and a detailed description of methods). **Figure S1** provides a full description of sample exclusions and missing data. Exclusion criteria for this study included prior serious medical and/or neurological illnesses unrelated to TBI, history of moderate to severe TBI, active suicidal and/or homicidal ideation requiring intervention, current diagnosis of bipolar disorder or psychotic disorder (except psychosis not otherwise specified due to trauma-related hallucinations), and exaggerated symptoms (score ≧ 23) on the Neurobehavioral Symptom Inventory. The study was approved by the VA Boston Healthcare System Institutional Review Board and all participants provided written informed consent.

### Clinical measures

#### Demographics, blast exposure, & mTBI

A demographics questionnaire was used to collect sex, ethnicity, education, and health history.

The Boston Assessment of TBI-Lifetime (BAT-L; [21]), was used to assess participants’ history of blast exposure and mild, moderate, and severe TBI. The BAT-L is a validated, semi-structured clinical interview administered by a doctoral-level psychologist that is used to assess a lifetime history of TBI and blast exposure. It establishes a TBI diagnosis based on time estimates of loss-of-consciousness, posttraumatic amnesia, and altered mental status duration. In addition, the BAT-L assesses the number of military-related bast exposures that occurred within 0 - 10 meters (close blast) or 11 - 100 meters (mid-to-far range blast). Participants who reported blast exposure may have also reported an mTBI for the same incident; however, blast exposure is a measure of mere exposure rather than TBI symptomatology. For primary analyses, “blast” was defined as any exposure to close or mid-to-far range blast. “TBI” was defined as lifetime history of mTBI. For exploratory analyses, participants were divided into groups related to proximity of blast exposure (close and mid-to-far) and mTBI mechanism (blast and blunt). These groups are further defined below in Statistical Analysis.

### Psychiatric assessments

A doctoral-level psychologist conducted all psychiatric assessments. History of PTSD was assessed using the Clinician Administered PTSD Scale for the DSM-IV (CAPS; [22]). The CAPS-IV includes three symptom clusters: re-experiencing, avoidance/emotional numbing, and hyperarousal; however, previous research suggests a four-factor model fits PTSD symptomatology more accurately [23,24]. Therefore, we computed subscales based on a four-factor model of PTSD [25]. These subscales are re-experiencing, avoidance, dysphoria, and hyperarousal. The subscales had excellent reliability in our sample (α ranged from .85 to .92).

The Structured Clinical Interview for DSM-IV Axis I Disorders (SCID; [25–27]) was used to screen for psychotic disorders for exclusionary criteria. Each week, the BAT-L, CAPS diagnoses, and SCID were reviewed by at least three doctoral level psychologists for diagnostic consensus.

### Pain

The Short Form McGill Pain Questionnaire (SF-MPQ; [28,29]) was used to assess pain. The SF-MPQ consists of 15 sensory and affective descriptors of pain that are rated on a scale from 0 (None) to 3 (Severe). These pain descriptor ratings are summed to create a pain index rating that ranges from 0 to 45. The SF-MPQ had excellent reliability in our sample (α = .94).

### Neurobehavioral symptoms

Neurobehavioral symptoms were indexed by the Neurobehavioral Symptom Inventory (NSI; [30]). The NSI consists of four subscales that measure vestibular, somatic, cognitive, and affective domains [31]. The NSI is a 22-item questionnaire, each item is rated from 0 (None) to 5 (Very Severe), and the NSI Total Score is obtained by summing scores across items. Due to the generalized nature of the symptoms queried (e.g., headaches, sleep difficulty, concentration problems), the NSI has been shown to be sensitive to common mental health comorbidities such as PTSD in addition to mTBI in military/Veteran populations [32–34]. NSI full-scale reliability was excellent in our sample (α = .97) and the subscale reliability ranged from good to excellent in our sample (α ranged from .85 to .91).

### Depression, anxiety, and stress

Participants were administered the Depression Anxiety and Stress Scale (DASS; [35]). The DASS is a 21 item self-report questionnaire that consists of seven questions for each category: depression, anxiety, and stress. Each question is rated by the respondent on a scale of 0 (did not apply to me at all) to 3 (applied to me very much, or most of the time). Scores for items within each category are summed to provide category subscales. In our sample, the DASS full scale had good reliability (Cronbach’s α =. 95) and the subscale reliability ranged from good to excellent in our sample (α ranged from .83 to .92).

### Biomedical measures

#### Simoa plasma assays

Participants were asked to fast before arriving to the study site. Informed consent was obtained between 7:00am and 7:30am and blood was drawn immediately thereafter. Plasma was collected in ethylenediaminetetraacetic acid (EDTA) tubes and centrifuged before being frozen and stored at −70 degrees until assayed. Simoa assays were conducted in batches to avoid multiple freeze-thaw cycles. Assays to measure plasma biomarker concentrations were run in duplicate using the ultra-sensitive single molecule array (Simoa technology developed by Quanterix, Billerica, MA). The samples were aliquoted into 96 well plates using the same template for each plate and were diluted to the manufactures’ specifications. For each kit, samples were run in duplicate. The lab technician was blinded to the clinical status of participants. The assays were run on the automated high-definition 1 (HD-1) analyzer. Plasma samples were not used if the reported coefficients of variation (CV) were over 20% and below the lower limit of quantification (LLOQ). The average CV was 5.10% for IL-6 and 5.15% for TNF-α. The LLOQ was 0.011 for IL-6 and 0.051 for TNF-α. To correct for non-normality, inflammation marker data were log transformed.

### Metabolic symptoms and serum assays

Metabolic data were collected by a medical technician and included blood pressure, height, weight, waist-to-hip ratio, body mass index, and a blood draw. Mean arterial blood pressure was a composite variable computed from two averaged systolic and one diastolic blood pressure measurements. Blood samples for CRP, fasting glucose, HDL, triglycerides, and A_1C_ were processed immediately and shipped on the same day to a commercial lab for metabolic panels. The assay for CRP used monoclonal antibodies with an analytical sensitivity of 0.10 mg/dL. Data for CRP was log transformed to correct for non-normality.

### Statistical analysis

#### Transdiagnostic symptom latent model & inflammation composite

CFA using Lavaan package in R version 4.0.2 [36] was conducted to examine the acceptability of a four-factor model of deployment-related transdiagnostic symptoms. CFA is used to produce theoretically meaningful latent constructs that capture the variance in common across measured variables. The four latent variables were “traumatic stress,” “affective,” “somatic,” and “metabolic.” Traumatic stress was defined by four CAPS subscales (re-experiencing, avoidance, dysphoria, and hyperarousal). Affective was defined by the DASS subscale scores (depression, anxiety, and stress), along with the NSI affective and cognitive subscales. Somatic was defined by NSI somatic and vestibular subscales and the SF-MPQ overall pain score. Finally, metabolic was defined by mean arterial blood pressure, hip to waist ratio, body mass index, fasting glucose, high-density lipoprotein, triglycerides, and hemoglobin A_1C_. Because our data were not normally distributed, we conducted the CFA using maximum likelihood estimation with robust standard errors. Four criteria were assessed for goodness of fit [37] - Comparative Fit Index (CFI; > 0.90), Tucker Lewis Index (TLI; > 0.90), Root Mean Square Error of Approximation (RMSEA; < 0.08), and Standardized Root Mean Square Residual (SRMR; < 0.06). Chi-square values were not used to assess goodness as they can be skewed by large sample sizes.

A composite variable for inflammation was computed from the average of the log transformed inflammation variables (IL-6, TNF-α, and CRP). To assess the relationships between the inflammatory markers and the composite variable, a partial correlation, adjusting for age, was conducted.

#### Blast/mTBI moderation analyses

Eight separate moderation models were conducted to examine the interaction between blast exposure or mTBI and each of the factors on the inflammation factor. In each model, age was entered in step 1. In step 2, either blast or mTBI was entered along with a latent variable, and in step 3, the interaction of blast or mTBI × latent variable was entered (e.g., blast × traumatic stress and mTBI × traumatic stress). The Benjamini-Hochberg False Discovery Rate (FDR) correction was applied to adjust for multiple comparisons. Moderation analyses were run with gender as a covariate to examine sex differences. Sex was not a significant variable in the analyses and the results did not change with or without it in the model. Therefore, we did not include it in the final models.

#### Blast & mTBI mechanism

Exploratory analysis of covariance (ANCOVA) was conducted to examine the relationship between the distance between the veteran and the blast detonation point, history of mTBI, and the metabolic factor, adjusting for age. Three groups were categorized by blast distance on the metabolic factor: 1) no blast exposure (*n* = 79), 2) blast exposure between 11 - 100 meters (mid-to-far range blast; *n* = 136), and 3) blast exposure between 0 - 10 meters (close blast; *n* = 142).

The relationship between mechanism of mTBI (blast vs. blunt) on the metabolic factor was examined by dividing participants into four groups, two without mTBI and two with mTBI: 1) control participants with no history of blast exposure or mTBI (*n* = 43), 2) participants with close blast exposure but no history of mTBI (*n* = 32), 3) participants with blunt-related mTBI but no history of blast exposure (*n* = 54), and 4) participants with blast-related mTBI but no history of blunt-related mTBI (*n* = 50).

## RESULTS

### Demographics and clinical characteristics

**Table 1** summarizes demographic and clinical characteristics of the sample (see **Table S1** for additional sample characteristics). Participants on average were 33 years old (SD = 8.9), mostly White (75.05%), and mostly men (90%). Just over half of the sample had current PTSD (58.54%) and had experienced at least one mTBI (66.95%). 77.87% had experienced at least one blast exposure at less than 100 meters. Five participants were diagnosed with diabetes (Type 1 = 3; Type 2 = 2). These participants were not outliers on any of the variables examined. **Table S2** reports clinical characteristics by blast and mTBI groups.

**Table 1.** Demographics and Clinical Characteristics.

Demographics and Clinical Characteristics
|  | *n or M* | *% or SD* |
| --- | --- | --- |
| Age | 33.04 | 8.91 |
| Men | 320 | 89.60% |
| **Race** |  |  |
| White, Non-Hispanic | 268 | 75.05% |
| Black, Non-Hispanic | 28 | 7.84% |
| Hispanic | 51 | 14.28% |
| Native American | 4 | 1.12% |
| Asian | 5 | 1.40% |
| Pacific Islander | 1 | 0.28% |
| **Blast Exposure** |  |  |
| 0 - 10 Meters | 3.17 | 24.26 |
| 11 - 100 Meters | 21.20 | 80.80 |
| **Traumatic Brain Injury** |  |  |
| Lifetime TBI | 239 | 66.95% |
| Blast-Related TBI | 95 | 26.61% |
| Years Since TBI | 9.44 | 8.39 |
| **Clinical Comorbidities** |  |  |
| PTSD | 209 | 58.54% |
| Current Mood | 89 | 24.93% |
| Current Anxiety | 63 | 17.65% |
| Current Substance Use | 59 | 16.53% |

*Note.* $n = 357$ . Blast exposure proximity refers to the mean number and standard deviation (SD) of blast exposures encountered at either 0-10 meters or 11-100 meters. Years since TBI refers to the mean number of years and SD since the last TBI was experienced.

### Transdiagnostic symptom latent model

The CFA of transdiagnostic health symptoms demonstrated good fit to the four-factor model reflecting traumatic stress, affective, somatic, and metabolic latent variables (CFI = 0.96, TLI = 0.95, SRMR = 0.04, and RMSEA = 0.52). Each latent variable showed significant associations with their respective indicators at *p* < .001 and standardized βs ranged from -.471 to .937. **Figure 1** provides pathway β loadings for each latent variable. **Table S3** provides loadings for each indicator on the latent variable.

**Figure 1.**
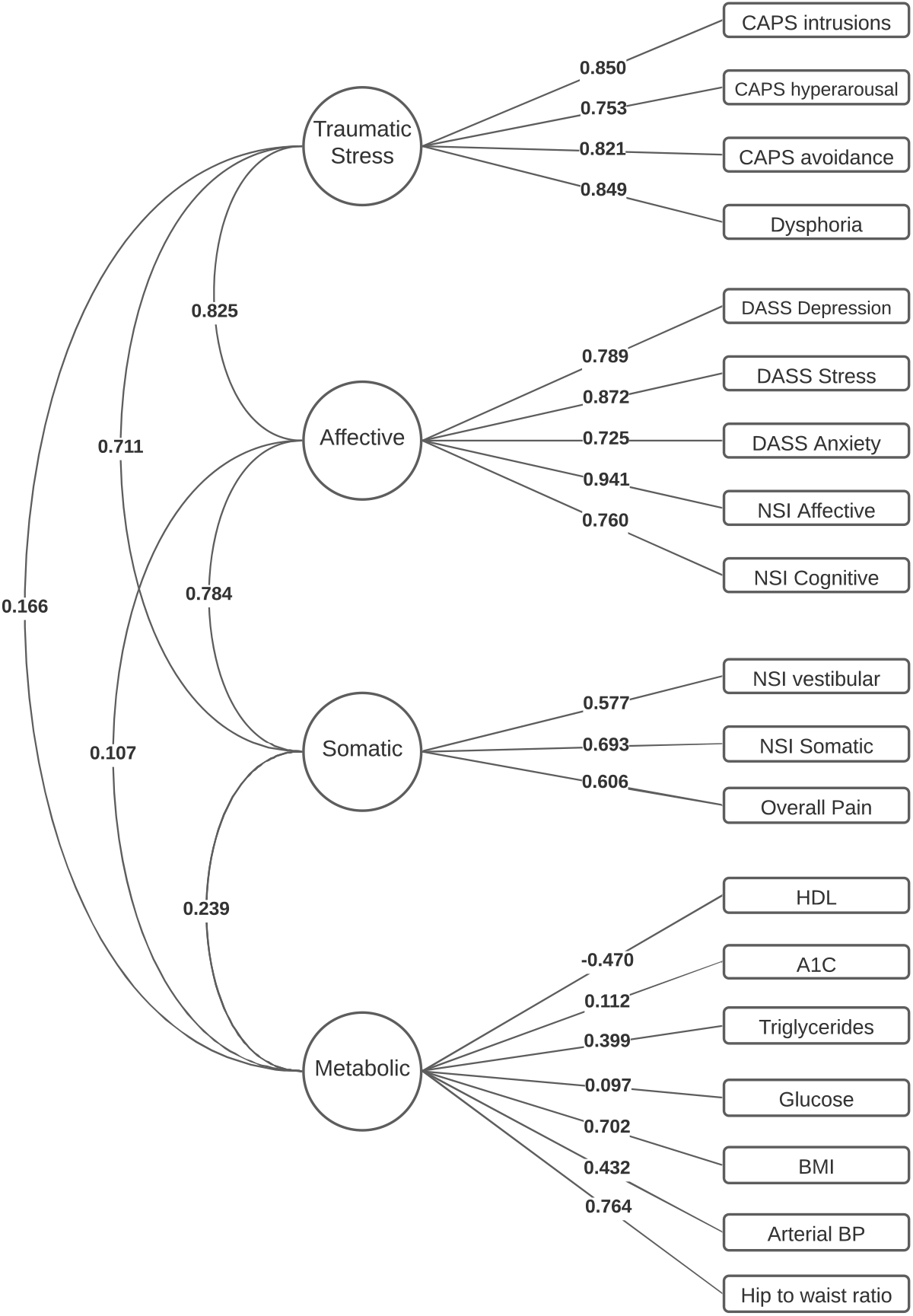
Transdiagnostic Symptom Latent Model.

### Inflammation composite

Results showed that each of the inflammatory markers were strongly associated with the overall inflammation composite (Log transformed IL-6, *r*[448] = .818, *p* < .001; TNF-α, *r*[448] = .615, *p* < .001; and CRP, *r*[448]= .852, *p* < .001). Thus, the inflammation composite was used as the outcome variable for all analyses.

### Blast/mTBI, latent variables, & inflammation Traumatic stress factor

Hierarchical regression analysis revealed that traumatic stress symptom severity was associated with higher levels of inflammation (β = .144, *p* = .009), consistent with prior literature linking PTSD symptoms to inflammation (see **Table 2**). However, the blast × traumatic stress interaction (β = -.066, *p* =.604) was not significant.

**Table 2.**
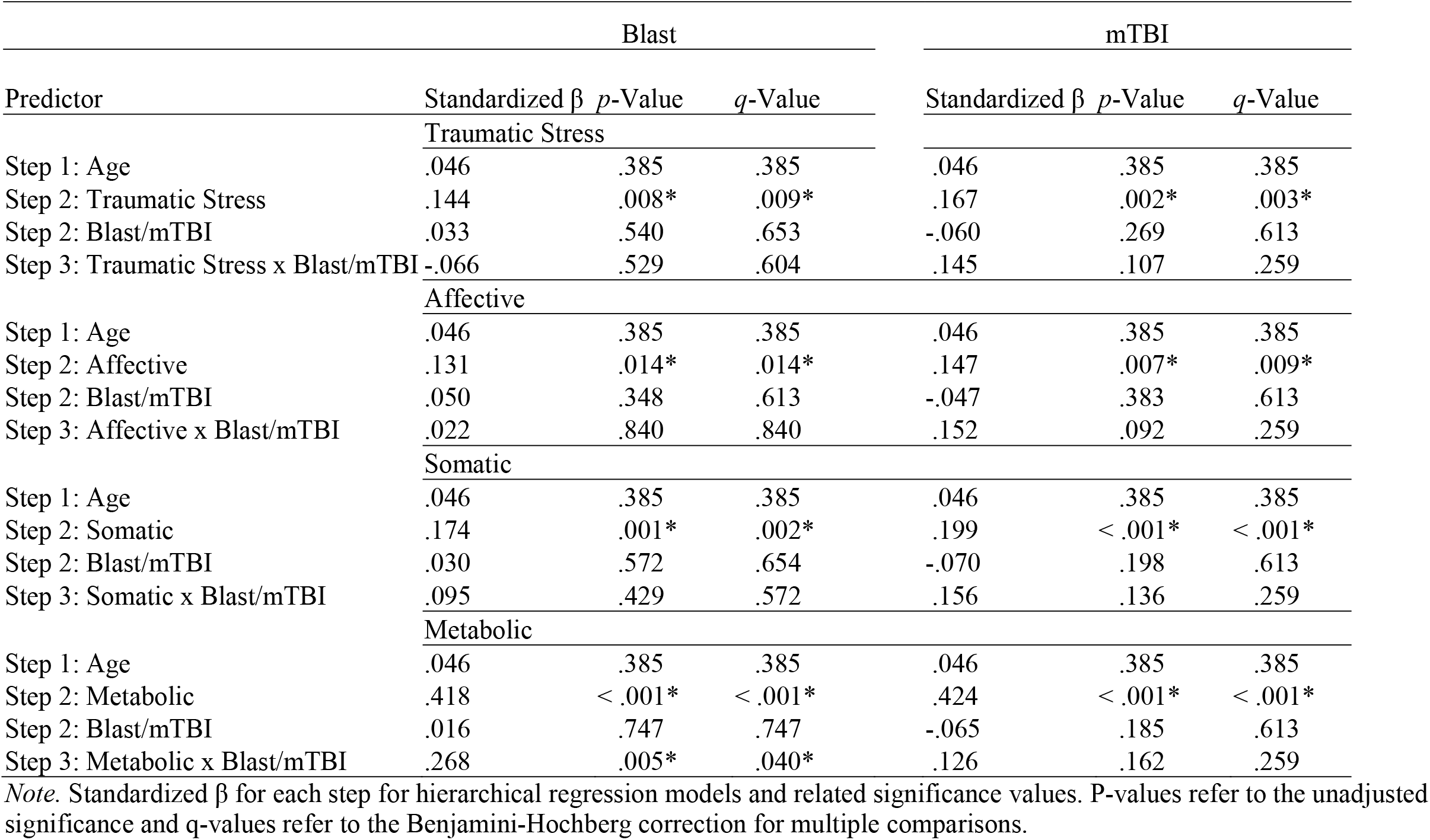
Hierarchical Regression Analysis Standardized β loadings.

| Predictor | Blast |  |  | mTBI |  |  |
| --- | --- | --- | --- | --- | --- | --- |
| | Standardized $\beta$ | *p*-Value | *q*-Value | Standardized $\beta$ | *p*-Value | *q*-Value |
| Traumatic Stress |  |  |  |  |  |  |
| Step 1: Age | .046 | .385 | .385 | .046 | .385 | .385 |
| Step 2: Traumatic Stress | .144 | .008* | .009* | .167 | .002* | .003* |
| Step 2: Blast/mTBI | .033 | .540 | .653 | -.060 | .269 | .613 |
| Step 3: Traumatic Stress x Blast/mTBI | -.066 | .529 | .604 | .145 | .107 | .259 |
| Affective |  |  |  |  |  |  |
| Step 1: Age | .046 | .385 | .385 | .046 | .385 | .385 |
| Step 2: Affective | .131 | .014* | .014* | .147 | .007* | .009* |
| Step 2: Blast/mTBI | .050 | .348 | .613 | -.047 | .383 | .613 |
| Step 3: Affective x Blast/mTBI | .022 | .840 | .840 | .152 | .092 | .259 |
| Somatic |  |  |  |  |  |  |
| Step 1: Age | .046 | .385 | .385 | .046 | .385 | .385 |
| Step 2: Somatic | .174 | .001* | .002* | .199 | < .001* | < .001* |
| Step 2: Blast/mTBI | .030 | .572 | .654 | -.070 | .198 | .613 |
| Step 3: Somatic x Blast/mTBI | .095 | .429 | .572 | .156 | .136 | .259 |
| Metabolic |  |  |  |  |  |  |
| Step 1: Age | .046 | .385 | .385 | .046 | .385 | .385 |
| Step 2: Metabolic | .418 | < .001* | < .001* | .424 | < .001* | < .001* |
| Step 2: Blast/mTBI | .016 | .747 | .747 | -.065 | .185 | .613 |
| Step 3: Metabolic x Blast/mTBI | .268 | .005* | .040* | .126 | .162 | .259 |

*Note.* Standardized $\beta$ for each step for hierarchical regression models and related significance values. P-values refer to the unadjusted significance and q-values refer to the Benjamini-Hochberg correction for multiple comparisons.

When considering mTBI instead of blast in the regression model, traumatic stress was associated with higher inflammation (β = .167, *p* = .003) but not the mTBI × traumatic stress interaction (β = .145, *p* = .259).

### Affective factor

Hierarchical models revealed that affective symptom severity was associated with higher inflammation (β = .131, *p* = .014), consistent with prior literature linking depression and anxiety symptoms to inflammation. However, the blast × affective and the mTBI × affective interaction were not significant predictors of inflammation.

### Somatic factor

Regression analysis revealed that somatic symptom severity was associated with higher inflammation (β = .174, *p* = .002). These results provide evidence of somatic indicators such as pain and vestibular dysregulation are associated with higher inflammation. However, the blast × somatic and mTBI × somatic interaction were not significant.

### Metabolic factor

When examining the metabolic factor in relation to blast and inflammation, regression analyses revealed a significant blast × metabolic factor interaction (*R*^*2*^ = .182, *F*[4, 352] = 19.54, *p* < .001). By contrast, the mTBI × metabolic interaction was not significant.

To parse the blast × metabolic interaction, we divided metabolic severity scores by median split. In the high metabolic group, blast exposure was associated with elevated inflammation *F*(2, 175) = 4.09, *p* = .045. However, there was no significant difference in inflammation between those with and without blast exposure in the low metabolic symptom group *F*(2, 176) = 0.66, *p* = .418. These results suggest that individuals with blast in the context of high metabolic symptoms had higher inflammation levels than those with low metabolic symptoms (see **Figure 2**).

**Figure 2.**
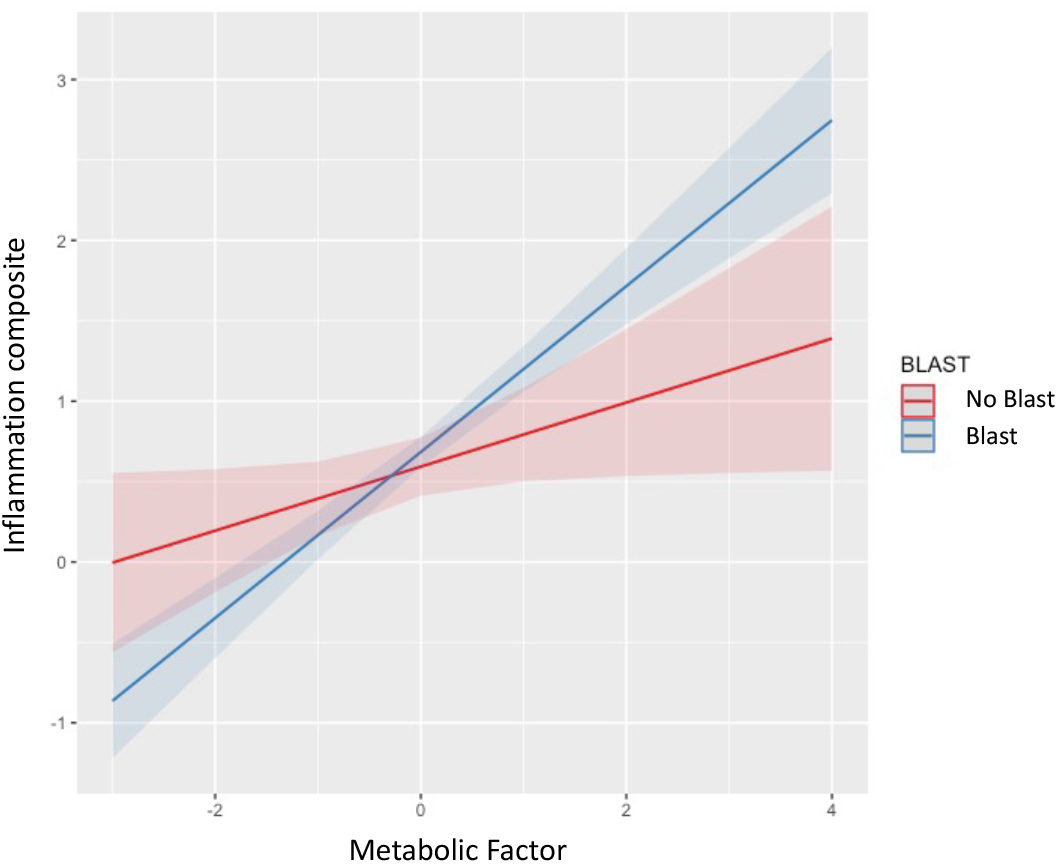
Blast Moderates the Relationship between Metabolic Factor and Inflammation.

### Sensitivity analysis

To determine the blast × metabolic effects on inflammation after taking into account the traumatic stress, affective, and somatic latent variables, hierarchical regression was conducted including all of the latent variables in a single model. Results revealed that the blast × metabolic interaction remained a significant predictor of inflammation (*R*^*2*^ = .191, *F*[4, 352] = 11.74, *p* < .001), even after adjusting for the other latent variables (see **Table S4**).

### Exploring the relationship between blast exposure and metabolic symptoms

The results from the primary analyses revealed a specific relationship between blast exposure and inflammation as a function of metabolic symptom severity. To further understand these findings, we explored the hypothesis that closer blast proximity would relate to greater metabolic symptom severity. Participants were divided into three groups: 1) no blast exposure; 2) blast exposure between 11 - 100 meters (mid-to-far blast); and 3) blast exposure between 0 - 10 meters (close blast). ANCOVA, adjusting for age and with metabolic factor score as the dependent variable, revealed a significant model *F*(2, 353) = 9.443, *p* < .001, η_p_^2^ = .051. Pairwise comparisons, after applying the Benjamini-Hochberg correction for multiple comparisons, revealed that individuals with close blast had significantly higher metabolic factor scores compared to individuals with mid-to-far range blast (*p* = .031) and those with no blast exposure (*p* < .001). Individuals with mid-to-far range blast exposure had higher metabolic factor scores than those with no blast exposure (*p* = .023). Together, these findings indicate a dose-response relationship such that metabolic symptoms are more severe in those individuals who are close to the blast.

To examine the effect of TBI mechanism (blast vs blunt) on metabolic symptom severity, we divided participants into four groups: 1) no history of blast exposure or mTBI; 2) close blast exposure but no history of mTBI; 3) blunt-related mTBI but no history of blast exposure; and 4) blast-related mTBI but no history of blunt-related mTBI. ANCOVA showed a significant model, after adjusting for age, *F*(3, 133) = 5.78, *p* = .001, η_p_^2^ = .115. Pairwise comparisons, after applying the Benjamini-Hochberg correction for multiple comparisons, revealed that individuals with blast-related mTBI had significantly higher metabolic factor scores compared to individuals with blunt mTBI (*p* = .004), and those with no blast exposure (*p* < .001). Individuals with close blast but no history of mTBI had significantly higher metabolic factor scores than those with no blast exposure (*p* = .047). **Figure 3** displays ANCOVA results for both models. These results suggest that blast mechanism, but not blunt injury, is associated with metabolic dysregulation.

**Figure 3.**
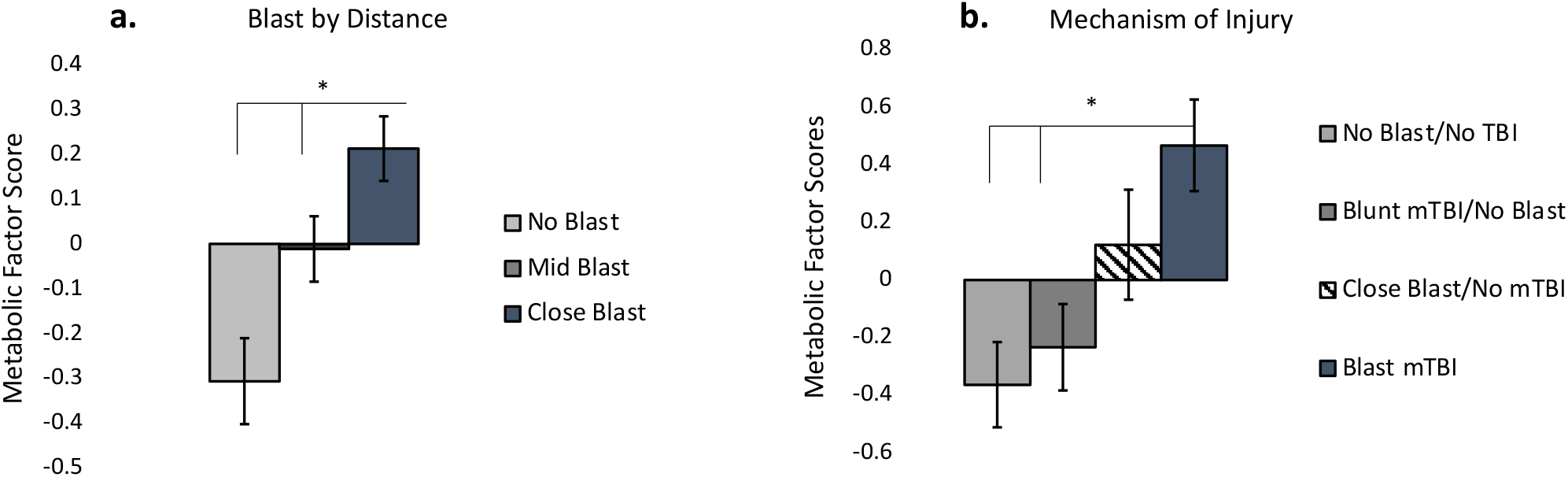
Relationships between Metabolic Factor, Blast distance, and mechanism of injury.

## DISCUSSION

The goal of this study was to determine the relationships between blast exposure, mTBI, transdiagnostic health symptoms, and systemic inflammation. There were three main findings. First, transdiagnostic mental and physical health symptoms (traumatic stress, affective, somatic, metabolic latent variables) extracted from a rich dataset of health variables were each associated with higher levels of systemic inflammation. However, the strongest relationship with inflammation emerged among those who had both war-zone blast exposures and metabolic dysregulation, even after adjusting for mental health latent variables. Finally, exploratory analyses showed that blast exposure was associated with metabolic dysregulation in a dose-response manner, with closer proximity to the blast associated with the greatest metabolic dysregulation. Together, these results provide greater understanding of the types of symptoms most strongly associated with inflammation, and the health pathways that link blast injury to systemic inflammation.

Chronic systemic inflammation is of great concern for a variety of reasons, including long-term negative health outcomes such as neurodegenerative disease. Biomarker studies have separately linked PTSD (reviewed in [38]), depression [39], and anxiety [40] to pro-inflammatory cytokine levels. Moreover, somatic symptoms are associated with higher CRP levels [41]. Across disorders that share these symptoms, there is evidence for hypothalamic-pituitary-adrenal (HPA) axis dysregulation in response to stress, which may lead to greater release of inflammatory cytokines. While the results reported here support the transdiagnostic nature of inflammation, they also demonstrate for the first time that the combination of blast injury and metabolic dysregulation is of particular significance for systemic inflammation, even after adjusting for traumatic stress, affective, and somatic symptoms. We did not find a direct relationship between blast exposure and mTBI with elevated inflammation, or an interaction of blast/mTBI with the mental health latent variables, despite some research demonstrating a direct link [9]. Rather, the association between blast exposure and inflammation differed as a function of metabolic symptom severity. These results are consistent with prior research showing a close relationship between blast and metabolic dysfunction [42].

Decades of work has demonstrated the association between core features of metabolic syndrome (e.g., obesity and insulin resistance) and abnormal inflammatory pathways (reviewed in [19]). Relevant here, there is growing evidence linking blast exposure to metabolic and endocrine abnormalities. While the mechanisms are not well understood, hypothalamic dysregulation and hypopituitarism are likely candidates, along with HPA axis and sympatho-adrenal medullary axis alterations [43]. Following blast injury, orexin A, an amino acid that regulates endocrine function of the hypothalamus, is elevated [44] and may explain how blast induces hypertension and the development of obesity [45]. Furthermore, blast induces hormonal changes associated with hypothalamic dysfunction and changes in the distribution of adipose tissue [42]. These data provide a potential mechanism for the link between blast injury and metabolic conditions, such as obesity, observed in the Veteran population [46] and support the link between blast and the metabolic latent variable observed in the present study.

In US service members, blast injury is associated with higher rates of hypopituitarism than those without such exposure [47]. Other work has shown that pre-existing metabolic conditions exacerbate inflammatory levels following TBI. In one study, animals fed high-fat diets had greater microglial activation and worse cognitive function following TBI [48]. These data suggest that obesity-related metabolic changes reduce the body’s capacity for repair following head injury, and are consistent with our findings showing greater metabolic dysregulation the in the blast exposed TBI group.

Interestingly, we did not find a relationship between blunt injury and metabolic syndrome, but rather blast exposures, inconsistent with the work showing metabolic changes in sports concussion [49]. It may be that the blunt injuries in this study were not of sufficient severity to observe metabolic imbalances seen in repetitive injuries sustained in professional sports. Notwithstanding, our exploratory findings suggest that blast exposures in and of themselves are related to metabolic dysfunction.

Limitations of the present study should be noted. Because the relationships observed are cross-sectional, the direction of effects cannot be determined. For instance, some work has shown that inflammation may not be a consequence of mental health disorders, but rather drives the development of mental health symptoms. We also cannot determine whether the blast by metabolic interaction was driven by blast-induced metabolic changes, or whether blast injury exacerbated the effect of existing metabolic dysregulation. Longitudinal studies are necessary to parse out directional and temporal effects of blast, health variables and inflammation. Another limitation was that sex effects could not be fully examined given the smaller number of women in the study. Although we did not find evidence for sex effects, a larger study with more female Veterans is necessary to understand potential differences in health variables and systemic inflammation.

## Conclusion

We report here that war-zone blast exposure in the context of metabolic symptom severity (e.g., high BMI, waist-to-hip ratio, hypertension) is associated with higher levels of the systemic inflammatory markers IL-6, TNF-a, and CRP. These results contribute to our current understanding of inflammation by underscoring the importance of metabolic symptoms relative to other types of symptoms in elevating inflammatory markers. An important takeaway from the these results is that particular emphasis should be placed on maintaining a healthy lifestyle to reduce the impact of obesity and other metabolic symptoms on future chronic disease, alongside TBI and blast prevention and psychiatric treatment for traumatic stress, depression, and anxiety in younger to middle-aged Veterans.

## Data Availability

The data are not publicly available.

## FUNDING & DISCLOSURE

This work was supported by the National Institutes of Health (NIH), NIA grant number R01AG058822 to JPH. The content is solely the responsibility of the authors and does not necessarily represent the official views of the NIH. This research was also supported in part by the Department of Veterans Affairs by the Translational Research Center for TBI and Stress Disorders (TRACTS), a VA Rehabilitation Research and Development Traumatic Brain Injury Center of Excellence (B9254-C). The authors declare no conflict of interest.

## ACKNOWLEDGMENTS

We thank Dr. Paul De Boeck for his help on the project.

## AUTHOR CONTRIBUTIONS

JPH was the principal investigator for the study and drafted the initial manuscript with contributions from MP and KV. RM, WM, CF, and JF collected, processed, and managed the TRACTS data. BRH processed the Simoa biomarker data. JPH, MP, KV, MM, RM, and WM contributed to data analysis plan and interpretation. All authors contributed to the final manuscript.

## FIGURE LEGENDS

**Figure 1**. Confirmatory factor analysis model for four factors related to transdiagnostic symptoms in post-9/11 Veterans.

**Figure 2**. Blast exposure moderates the relationship between metabolic factor and inflammation.

**Figure 3**. a) Close range blast is related to greater metabolic symptom severity compared to mid-far ranged blasts and (*p* = .031) no blast exposure (*p* = .023). b) Blast-related mTBI is related to higher metabolic symptom severity compared to blunt-related mTBI (*p* = .004) and no mTBI (*p* < .001).

**Figure S1**. Exclusion Process and missing data.

## Notes

### Competing Interest Statement

The authors have declared no competing interest.

### Author Declarations

The study was approved by the VA Boston Healthcare System Institutional Review Board and all participants provided written informed consent.

