## Supplemental for "The association between blast exposure and transdiagnostic health symptoms on systemic inflammation"

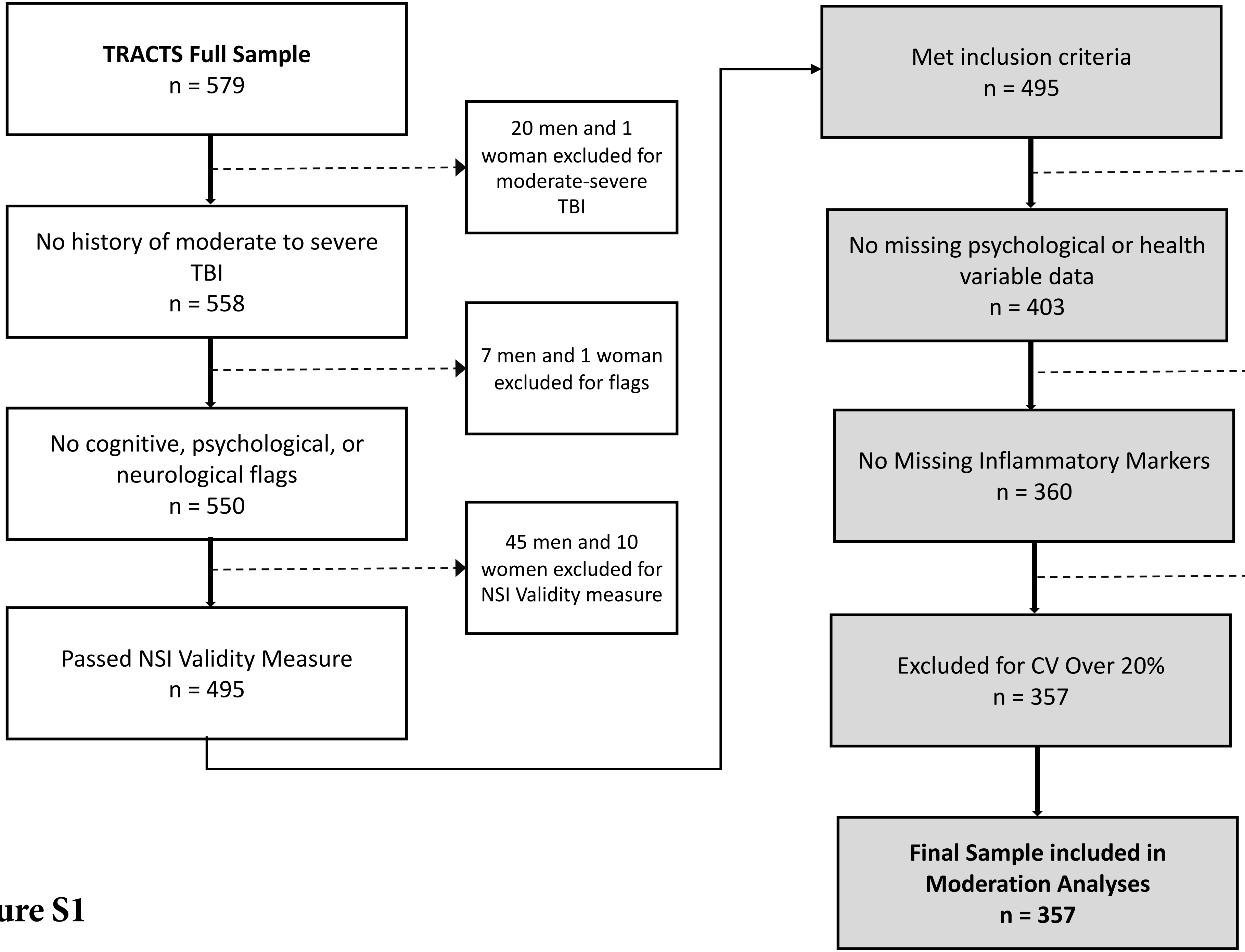
**TRACTS Full Sample**
n = 579
20 men and 1 woman excluded for moderate-severe TBI
No history of moderate to severe TBI
n = 558
7 men and 1 woman excluded for flags
No cognitive, psychological, or neurological flags
n = 550
45 men and 10 women excluded for NSI Validity measure
Passed NSI Validity Measure
n = 495
Met inclusion criteria
n = 495
86 men and 6 women with missing data
No missing psychological or health variable data
n = 403
37 men and 6 woman with missing data
No Missing Inflammatory Markers
n = 360
3 men and 0 women with CV over 20%
Excluded for CV Over 20%
n = 357
**Final Sample included in Moderation Analyses**
**n = 357**

**Figure S1**

**Table S1**

Demographic Information for Blast and mTBI Groups

| | No Blast Exposure n=79 | | Blast Exposed n=278 | | No History of mTBI n=118 | | History of mTBI n=239 | |
|---|---|---|---|---|---|---|---|---|
| | *n or M* | *% or SD* | *n or M* | *% or SD* | *n or M* | *% or SD* | *n or M* | *% or SD* |
| Age | 33.57 | 9.85 | 32.89 | 8.64 | 33.47 | 9.29 | 32.83 | 8.73 |
| Men | 64 | 81.01% | 255 | 91.73% | 100 | 84.75% | 220 | 92.05% |
| **Race** | | | | | | | | |
| White, Non-Hispanic | 55 | 69.62% | 213 | 76.62% | 76 | 64.41% | 192 | 80.34% |
| Black, Non-Hispanic | 8 | 10.13% | 20 | 7.19% | 12 | 10.16% | 16 | 6.69% |
| Hispanic | 11 | 13.92% | 40 | 14.39% | 28 | 23.73% | 23 | 9.62% |
| Native American | 2 | 2.53% | 2 | 0.72% | 1 | 0.85% | 3 | 1.25% |
| Asian | 3 | 3.80% | 2 | 0.72% | 1 | 0.85% | 4 | 1.68% |
| Pacific Islander | 0 | 0.00% | 1 | 0.36% | 0 | 0.00% | 1 | 0.42% |
| **Education** | | | | | | | | |
| High School or Less | 18 | 22.78% | 93 | 33.45% | 38 | 32.20% | 73 | 30.55% |
| Bachelor's Degree/Some College | 49 | 62.03% | 161 | 57.92% | 70 | 59.32% | 140 | 58.57% |
| Advanced Degree | 12 | 15.19% | 24 | 8.63% | 10 | 8.48% | 26 | 10.88% |
| **Service Branch** | | | | | | | | |
| Army | 48 | 60.76% | 189 | 67.99% | 83 | 70.34% | 154 | 64.44% |
| Navy | 6 | 7.59% | 13 | 4.67% | 4 | 3.39% | 15 | 6.27% |
| Marines | 11 | 13.92% | 63 | 22.66% | 19 | 16.10% | 55 | 23.01% |
| Airforce | 14 | 17.73% | 11 | 3.96% | 11 | 9.32% | 14 | 5.86% |
| Coast Guard | 0 | 0.00% | 2 | 0.72% | 1 | 0.85% | 1 | 0.42% |
| **Unit Type** | | | | | | | | |
| National Guard | 55 | 69.62% | 132 | 47.48% | 80 | 67.80% | 107 | 44.77% |
| Reserve | 24 | 30.38% | 146 | 52.52% | 38 | 32.20% | 132 | 55.23% |

**Table S2**

Clinical Characteristics for Blast and mTBI Groups

| | No Blast Exposure n=79 | | Blast Exposed n=278 | | No History of mTBI n=118 | | History of mTBI n=239 | |
|---|---|---|---|---|---|---|---|---|
| | *M or n* | *SD or %* | *M or n* | *SD or %* | *M or n* | *SD or %* | *M or n* | *SD or %* |
| **Blast Exposure** | | | | | | | | |
| 0 - 10 Meters | 0 | 0 | 4.06 | 27.44 | 0.44 | 1.14 | 4.51 | 29.57 |
| 11 - 100 Meters | 0 | 0 | 27.23 | 90.69 | 6.89 | 20.35 | 28.27 | 97.01 |
| **Traumatic Brain Injury** | | | | | | | | |
| Lifetime TBI | 0.89 | 1.39 | 1.74 | 2.33 | 0 | 0 | 2.32 | 2.32 |
| Blast-Related TBI | 0 | 0 | 0.44 | 0.74 | 0 | 0 | 0.52 | 0.79 |
| Years Since TBI | 13.95 | 11.28 | 8.56 | 7.43 | 0 | 0 | 9.44 | 8.39 |
| **Combat Exposure** | | | | | | | | |
| DRRI Combat | 6.36 | 5.95 | 19.96 | 10.87 | 12.27 | 9.37 | 19.55 | 11.62 |
| DRRI Other War Experiences | 3.99 | 3.91 | 9.27 | 4.43 | 5.38 | 4.17 | 9.53 | 4.52 |
| **Clinical Comorbidities** | | | | | | | | |
| *Diagnoses* | | | | | | | | |
| PTSD | 30 | 37.97% | 179 | 64.38% | 53 | 44.92% | 156 | 65.27% |
| Current Mood | 18 | 22.78% | 71 | 25.54% | 25 | 21.18% | 64 | 22.94% |
| Current Anxiety | 20 | 25.32% | 43 | 15.47% | 18 | 15.25% | 45 | 18.82% |
| Current Substance Use | 14 | 17.72% | 45 | 16.18% | 18 | 15.25% | 41 | 17.15% |
| *Symptom Severity* | | | | | | | | |
| CAPS Symptoms Severity | 34.66 | 32.06 | 51.90 | 28.18 | 37.18 | 30.25 | 53.47 | 28.28 |
| DASS Depression Severity | 7.59 | 8.92 | 8.89 | 9.49 | 7.14 | 9.49 | 9.33 | 9.25 |
| DASS Anxiety Severity | 4.94 | 6.99 | 7.11 | 7.33 | 4.86 | 6.28 | 7.50 | 7.62 |
| DASS Stress Severity | 11.29 | 10.93 | 13.56 | 9.78 | 10.54 | 10.59 | 14.30 | 9.59 |

*Note.* Blast exposure proximity refers to the mean number and standard deviation (SD) of blast exposures encountered at either 0-10 meters or 11-100 meters. Years since TBI refers to the mean number of years and SD since the last TBI was experienced.

**Table S3**

Latent Variable Loadings

| Latent Variable | Indicator | B | SE | Z | p-Value | β |
|---|---|---|---|---|---|---|
| Traumatic Stress | Intrusions | .850 | .031 | 27.68 | < .001 | .871 |
| Traumatic Stress | Avoidance | .821 | .028 | 28.98 | < .001 | .833 |
| Traumatic Stress | Hyperarousal | .753 | .033 | 22.66 | < .001 | .753 |
| Traumatic Stress | Dysphoria | .849 | .028 | 29.97 | < .001 | .865 |
| Affective | DASS Depression | .789 | .042 | 18.89 | < .001 | .823 |
| Affective | NSI Cognitive | .760 | .033 | 23.09 | < .001 | .809 |
| Affective | NSI Affective | .941 | .030 | 31.212 | < .001 | .955 |
| Affective | DASS Anxiety | .725 | .041 | 17.83 | < .001 | .803 |
| Affective | DASS Stress | .872 | .033 | 26.71 | < .001 | .891 |
| Somatic | NSI Vestibular | .577 | .040 | 14.26 | < .001 | .693 |
| Somatic | NSI Somatic | .693 | .039 | 17.630 | < .001 | .825 |
| Somatic | McGill Pain Overall | .606 | .046 | 13.16 | < .001 | .626 |
| Metabolic | BMI | .702 | .051 | 13.75 | < .001 | .713 |
| Metabolic | Waist to Hip Ratio | .764 | .050 | 15.41 | < .001 | .789 |
| Metabolic | Mean Arterial BP | .432 | .052 | 8.36 | < .001 | .437 |
| Metabolic | HDL Cholesterol | .470 | .051 | 9.29 | < .001 | .478 |
| Metabolic | Triglycerides | .399 | .052 | 7.68 | < .001 | .391 |
| Metabolic | Glucose | .097 | .029 | 3.32 | .001 | .180 |
| Metabolic | $A_{1C}$ | .112 | .026 | 4.28 | < .001 | .204 |

*Note.* Loading statistics for each indicator on the latent variable.

**Table S4**

Sensitivity analysis

| Predictor | $R^2_{adj}$ | $R^2/\Delta R^2$ | *F* | *p*-value |
|---|---|---|---|---|
| Step 1: Age | -.001 | .002 | 0.76 | .385 |
| Step 2: Traumatic Stress, Affective, Somatic, Metabolic, Blast | .157 | .171/.169 | 12.04 | < .001* |
| Step3: Blast x Metabolic | .174 | .191/.020 | 11.74 | < .001* |

| Predictor | Standardized β | *p*-value | *q*-value |
|---|---|---|---|
| Step 1: Age | .046 | .385 | .673 |
| Step 2: Traumatic Stress | -.055 | .591 | .800 |
| Step 2: Affective | .150 | .217 | .506 |
| Step 2: Somatic | -.026 | .800 | .800 |
| Step 2: Metabolic | .418 | < .001* | < .001* |
| Step 2: Blast | -.013 | .797 | .800 |
| Step 3: Blast x Metabolic | -.066 | .004* | .014* |

*Note*. Hierarchical regression results including adjustment for Traumatic Stress, Affective, and Somatic factors. P-values refer to the unadjusted significance and q-values refer to the Benjamini-Hochberg correction for multiple comparisons.
